# Genetic inhibition of angiopoietin-like protein-3, lipoprotein-lipid levels and cardiometabolic diseases

**DOI:** 10.1101/2023.05.29.23290580

**Authors:** Émilie Gobeil, Jérôme Bourgault, Patricia L. Mitchell, Ursula Houessou, Eloi Gagnon, Arnaud Girard, Audrey Paulin, Hasanga D. Manikpurage, Christian Couture, Simon Marceau, Yohan Bossé, Sébastien Thériault, Patrick Mathieu, Marie-Claude Vohl, André Tchernof, Benoit J. Arsenault

## Abstract

**Background:** RNA-based, antibody-based and gene editing-based therapies are currently under investigation to determine if the inhibition of angiopoietin-like protein-3 (ANGPTL3) could reduce lipoprotein-lipid levels and atherosclerotic cardiovascular diseases (ASCVD) risk. We used Mendelian randomization (MR) to determine whether genetic variations influencing ANGPTL3 liver gene expression, blood levels and protein structure could causally influence triglyceride and apolipoprotein B (apoB) levels as well as coronary artery disease (CAD), ischemic stroke (IS) and other cardiometabolic diseases.

**Methods:** We performed RNA-sequencing of 246 explanted liver samples to identify single-nucleotide polymorphisms (SNPs) associated with liver expression of *ANGPTL3*. We used genome-wide summary statistics of plasma protein levels of ANGPTL3 from the deCODE study (n=35,359). We also identified 647 carriers of *ANGPTL3* protein-truncating variants (PTVs) associated with lower plasma triglyceride levels in the UK Biobank. We performed two-sample MR using SNPs that influence *ANGPTPL3* liver expression or ANGPTPL3 plasma protein levels as exposure and cardiometabolic diseases as outcomes (CAD, IS, heart failure, non-alcoholic fatty liver disease, acute pancreatitis and type 2 diabetes). The impact of rare PTVs influencing plasma triglyceride levels on apoB levels and CAD was also investigated in the UK Biobank.

**Results:** In two-sample MR studies, common genetic variants influencing *ANGPTL3* hepatic or blood expression levels of ANGPTL3 had a very strong effect on plasma triglyceride levels, a more modest effect on LDL cholesterol, a weaker effect on apoB levels and no effect on CAD or other cardiometabolic diseases. In the UK Biobank, carriers of rare *ANGPTL3* PTVs providing lifelong reductions in median plasma triglyceride levels (-0.37 [interquartile range=0.41] mmol/L) had slightly lower apoB levels (-0.06±0.32] g/L) and similar CAD event rate compared to noncarriers (10.2% versus 10.9% in carriers versus noncarriers, p=0.60).

**Conclusions:** PTVs influencing ANGPTL3 protein structure as well as common genetic variants influencing *ANGPTL3* hepatic expression and/or blood protein levels exhibit a strong effect on circulating plasma triglyceride levels, a weak effect on circulating apoB levels and no effect on ASCVD. Near-complete inhibition of ANGPTL3 function in patients with very elevated apoB levels will likely be required to reduce ASCVD risk.

## Introduction

Elevated plasma lipoprotein and lipid levels including low-density lipoprotein (LDL) cholesterol and triglycerides are important risk factors for atherosclerotic cardiovascular diseases (ASCVD) and some metabolic diseases^1, 2^. Every lipoprotein that is positively associated with long-term ASCVD risk contains one apolipoprotein B (apoB) molecule. A strong and dose-dependent effect of apoB on ASCVD risk has been reported in multiple observational studies^3^. Results of several recent Mendelian randomization (MR) studies and randomized cardiovascular outcomes trials provided evidence that lipid-lowering therapies may be effective in reducing ASCVD risk if they provide significant reductions in plasma apoB levels regardless of their impact on other lipoprotein parameters such as triglycerides or high-density lipoprotein cholesterol levels^4–6^. Many triglyceride-lowering agents are currently under investigation for their potential cardiovascular benefits. While some triglyceride-lowering agents such as icosapent ethyl may provide cardiovascular benefits, other triglyceride-lowering agents such as fibrates or formulations of omega-3 fatty acids, may not provide cardiovascular benefits^7–9^ .

Plasma levels of angiopoietin-like protein-3 (ANGPTL3) play a critical role in triglyceride-rich lipoprotein metabolism^10, 11^. ANGPTL3 is mostly expressed in the liver. Rare and common genetic variations at the *ANGPTL3* locus are associated with lower plasma levels of LDL cholesterol and triglyceride levels^12, 13^. At least three ANGPTL3 inhibition strategies are currently under investigation for their impact on lipoprotein-lipid levels and potential cardiovascular benefits. Inhibition of ANGPTL3 synthesis by a liver-targeted antisense oligonucleotide has been shown to substantially reduce plasma triglyceride levels in patients with elevated triglyceride levels^14, 15^. Neutralization of the circulating ANGPTL3 protein by a monoclonal antibody has also been shown to reduce triglyceride levels and LDL cholesterol levels, respectively in patients with elevated triglyceride levels and homozygous familial hypercholesterolemia^16, 17^. A CRISPR-based therapy testing a one-time gene editing strategy intended to potently and durably reduce plasma triglyceride and LDL cholesterol levels by inactivating hepatic ANGPTL3 via the introduction of a premature stop codon in the *ANGPTL3* DNA sequence is also under development^18, 19^. Whether targeting ANGPTL3 via one of these strategies could ultimately prevent ASCVD events in high-risk patients remains to be demonstrated.

By establishing the impact of genetic variants predicting human traits on health outcomes (such as cardiometabolic traits and diseases), MR offers the possibility of investigating the association between genetically predicted drug targets and cardiometabolic outcomes in “natural” randomized clinical trial (as genetic variants are randomly acquired at meiosis and immune to reverse causation and confounding). One of the most replicated MR studies presented the impact of sequence variants at the *PCSK9* gene linked with low LDL-cholesterol levels and protection against coronary artery disease (CAD)^20^. Randomized clinical trials subsequently confirmed that PCSK9 neutralization with monoclonal antibodies prevents atherosclerosis progression and protects against short-term ASCVD events^21–23^. Previous genetic/MR studies testing the impact of rare and common variants at *ANGPTL3* have provided conflicting results. While rare variants at the *ANGPTL3* locus were associated with protection against CAD^24, 25^, more common variants linked with lifelong reductions in plasma triglyceride, LDL cholesterol and apoB levels were not associated with CAD^26^. In this study, only one independent genetic variant with an unknown function at the *ANGPTL3* locus was associated with triglyceride levels. Combining genomics with other OMIC strategies such as transcriptomics and proteomics offers the opportunity to better understand how the genetic regulation of *ANGPTL3* gene or protein expression might be related to cardiometabolic traits and diseases. These new developments could enable the investigation of the cardiometabolic impact of different ANGPTL3 inhibition strategies.

The first objective of this study was to identify genetic variants at the *ANGPTL3* locus influencing 1) liver *ANGPTL3* gene expression, 2) circulating ANGPTL3 protein levels and 3) ANGPTL3 protein structure to respectively mimic ANGPTL3 RNA-based therapies, antibodies and gene-editing strategies. We sought to determine whether genetic variations at the *ANGPTL3* locus influencing *ANGPTL3* gene expression, circulating concentrations of ANGPTL3 levels or rare ANGPTL3 sequence variants influence plasma lipoprotein-lipid levels as well as cardiometabolic diseases using a comprehensive MR approach. We also aimed at identifying and replicating genetic determinants of circulating ANGPTL3 levels.

## Methods

### Identification of genetic variants influencing liver ANGPTL3 expression

Patients undergoing bariatric surgery at the Quebec Heart and Lung Institute (*Institut universitaire de cardiologie et de pneumologie de Québec* [IUCPQ]) provided informed consent to participate to the institutional biobank (IUCPQ Obesity Biobank). Clinical information at the time of surgery (sex, age, anthropometry, medication use, medical history and comorbidities and glycemic, lipoprotein and liver enzyme profile) was available for all patients. A total of 246 participants passed genotyping and RNA sequencing quality controls. Baseline characteristics of the 246 participants of the cohort are presented in Supplementary Table 1. Liver samples were obtained by incisional biopsy of left lobe and were not cauterized. The grading and staging of histological lesions have been carried out according to the protocol of Brunt et al.^27^ by pathologists who were blind to the objectives of the study. The liver sample procedure and position of the liver sample are standardized for among all surgeons. A detailed description of DNA and RNA extraction, genome-wide genotyping, RNA sequencing, mapping of expression quantitative trait expression loci (eQTL) is available in Supplementary Methods.

### Genome-wide association studies of circulating ANGPTL3 levels

The instruments for genetically-predicted circulating ANGPTL3 protein levels were obtained from a genetic association study in the deCODE genetics cohort^28^. In this study, 4,719 plasma protein levels, including ANGPTL3, were measured in 35,559 Icelanders with 4,907 aptamers from SomaScan version 4. A detailed description of the proteomic measurements, the genotyping and genome-wide association analyses have been described previously^28^. For the replication, we used the Fenland population-based cohort, where Pietzner et al.^29^ performed genome-wide association study (GWAS) for 4775 plasma protein levels, including ANGPTL3, were measured in 10,708 European-descent participants using the SomaScan v4 assay.

### Genetic colocalization

Genetic colocalization at the *ANGPTL3* cis genetic region was performed to determine whether genes influencing ANGPTL3 plasma levels did so by acting in a tissue-specific manner. We used visceral (n=469) and subcutaneous (n=581) adipose tissues from the publicly available Genotype-Tissue Expression Project (GTEx, version 8) ^30^ and liver samples (n=246) from the IUCPQ Obesity Biobank. We employed genetic colocalization^31^ to test for a shared signal between the concentration of ANGPTL3 measured in plasma and the expression of the respective genes in these three tissues. Genetic association and colocalization of the loci significantly associated with ANGPTL3 blood levels in deCODE genetic and eQTLs were performed using the package COLOC^31^ with the default priors. Regions for testing were determined by dividing the *ANGPTL3* encoding gene region ±500 Kb into 0.1-cM chunks using recombination data. Evidence for colocalization was evaluated with the posterior probability for hypothesis 4 (PP.H4); there is an association for both traits and they are driven by the same causal variant. We deemed a PP.H4>0.80 as evidence for a highly likely shared signal between cis-eQTL/cis-pQTL pairing for a given tissue and a PP.H4>0.50 as evidence for a likely shared signal.

### Genome-wide association studies used for Mendelian randomization

The GWAS for lipoprotein/lipid levels, and cardiovascular/metabolic diseases, all publicly available, were selected on greatest expected statistical power (sample size and cases/controls ratio). We used apoB levels measured in 441,016 participants from the UK Biobank (UKB) ^5^ and triglyceride, LDL-C, HDL-C levels measured in 1,320,016 participants from the Global Lipids Genetics Consortium (GLGC) ^32^. For cardiovascular diseases, the recent GWAS meta-analysis of coronary artery disease from the CARDIoGRAMplusC4D and UKB (181,521 cases and 984,199 controls) ^33^, the ischemic stroke GWAS of MEGASTROKE and FinnGen (34,217 cases and 406,111 controls) ^34^, and the heart failure GWAS from HERMES and UKB (47,309 cases and 930,014 controls) ^35^ were used. For metabolic diseases, GWAS meta-analysis studies of acute pancreatitis from UKB, FinnGen, Estonian biobank (10,630 cases and 844,679 controls) ^36^, of chronic kidney disease from CKDGen and UKB (41,395 cases and 439,303 controls) ^37^, of non-alcoholic fatty liver disease from Estonian Biobank, deCODE genetics, UKB, INSPIRE+HerediGene (USA), FinnGen and eMERGE Network (16,532 cases and 1,240,188 controls) ^38^ ^39^, and of type 2 diabetes from DIAGRAM and UKB (74,124 cases and 824,006 controls) ^40^ were used. Relevant information on the GWAS summary statistics used throughout this study is presented in Supplementary Table 2.

### Mendelian randomization of the health impact of liver expression and protein levels of ANGPTL3

In MR, genetic variants are used as ‘instrumental variables’ (IVs) for assessing the causal effect of exposure ^41^ (here *ANGPTL3* gene hepatic expression or blood protein levels) on the outcome (here lipoprotein/lipid levels or cardiometabolic diseases). For the single instrument MR analysis, we selected the strongest cis-acting eQTL within 500 Kb on either side of the *ANGPTL3* gene region. We used the same strategy to identify the strongest cis-acting plasma protein quantitative trait locus (pQTL) associated with circulating ANGPTL3 protein levels from deCODE genetics. We obtained eQTL-specific and pQTL-specific Wald ratio estimates for *ANGPTL3* liver expression and ANGPTL3 blood levels on 11 outcomes. This estimate is obtained by dividing the estimated effect of the genetic instrument on the outcome by the estimated effect of the genetic instrument on the exposure. The Wald ratio estimates correspond to the log-transformed odds ratio (for a binary outcome) or to the SD (for a continuous outcome) change per unit change of the exposure^42^. Hypothesis Prioritization in multi-trait Colocalization (HyPrColoc)^43^ was used to test for a shared signal between *ANGPTL3* liver expression with triglycerides, apoB and CAD. We repeated the analysis, but this time with ANGPTL3 protein blood expression, levels of triglycerides and apoB as well as CAD. For the multi-cis MR analysis, we obtained pQTLs from deCODE. We retained SNPs that were strongly associated with ANGPTL3 protein blood levels using a p-value threshold ≤ 5 x 10^-8^, including only cis-acting pQTLs (chromosome 1, position: [62,597,520 - 62,606,313] ± 500 Kb). We then performed LD clumping for the instruments with the ieugwasr R package and the European 1000-genome LD reference panel to select only independent pQTLs (R^2^ < 0.1) in the *ANGPTL3* gene region. We restricted the instruments to variants with a MAF > 1%. We annotated variants using the Ensembl Variant Effect Predictor (VEP). Our instruments did not include any variants in the coding region of *ANGPTL3*, which prevented any potential epitope-binding artifacts. The strength of every instrument was evaluated with the Cragg-Donald F-statistic ^44^. The selected genetic variants (Supplementary Table 3), as indicated above, and their summary statistics allow the causal effect of each protein in the cohort to be assessed using the inverse variance weighted (IVW) estimator. Briefly, this approach combines the Wald ratio estimates across all instruments. IVW-MR is equivalent to a weighted linear regression of SNP-outcome associations on SNP-exposure associations where the contribution of each SNP to the overall effect is weighted by the inverse of the variance of the SNP-outcome association and with the intercept constrained to zero^41^.

### Mendelian randomization on the impact of triglyceride levels on apolipoprotein B levels

The causal effect of plasma triglyceride levels on apoB circulating levels was obtained with an univariable MR analysis using the summary statistics of two GWAS of apoB and triglyceride levels measured in 441,016 participants from the UK Biobank ^5^. As IVs, we selected genetic variants significantly associated with plasma triglyceride levels (p≤5×10^-8^) and independent of each other (LD R^2^ < 0.001 within a window of 1000 Kb). The causal impact of triglyceride-reducing SNPs on apolipoproteinB (apoB) levels was assessed using the IVW-MR method.

### Identification of protein-truncating variants in ANGPLT3 and PCSK9 in participants of the UK Biobank

The study design and population of the UKB have previously been reported^45^. UKB includes more than 500,000 participants between 40 and 69 years old who were recruited from the United Kingdom’s National Health Service (NHS) central registers between 2006 and 2010. Assessment for diseases and serological tests for these participants were done in 22 assessment centers during the same period. Data collection included a self-report questionnaire, physical measures, and blood, urine, and saliva sample collection. Analyses in UKB were conducted under the data application number 25205. UKB participants provided signed and informed consent for their participation in these respective studies. UKB has been approved by the North West Multi-Center Research Committee (MREC). A total of 407,668 White British UK Biobank participants were included in the current analysis. PTVs in *ANGPTL3* and *PCSK9* (the latter used as a positive control) were extracted from the 407,668 White British participants with available genotyping data for the corresponding gene in the UK Biobank and were identified from whole-exome sequencing, as previously described^46^. We selected those for whom triglyceride levels (field number 30870) and apoB levels (field number 30640) were available. PTVs were defined as variants causing the loss of an exon or a frameshift in the translation or if they introduced a stop codon, a splice acceptor or a splice donor site^47^. In order to confirm that the identified PTVs are considered as strong, we used the Ensembl variant effect predictor (VEP) tool^48^ to extract the Combined Annotation Dependent Depletion (CADD) Phred scores for each variant. We also used GnomAD^49^ to extract the predicted loss-of-function (pLoF) value for each PTV. We considered a PTV to be strong if it had either a CADD Phred score above 20 (in the top 1% most deleterious substitutions of the genome) or a high-confidence pLoF value. Noncarriers were defined as having none of the variants identified. This procedure was performed independently for *ANGPTL3* and *PCSK9*.

### Health impact of protein-truncating variants in ANGPLT3

For each PTVs in *ANGPTL3*, we performed a linear regression on circulating triglyceride levels between noncarriers and carriers. Health effects of PTVs were investigated if PTVs were significantly associated with a reduction of triglyceride levels in both analyses (p<0.05). All strong *ANGPTL3* PTVs carriers were then grouped together and the impact of carrier status on CAD was determined. To define prevalent CAD, we selected participants with ICD-10 codes for MI (I21.X, I22.X, I23.X, I24.1, or I25.2), other acute ischemic heart diseases (IHD) (I24.0, I24.8-9) or chronic IHD (I25.0-25.1, I25.5-25.9). In order to investigate the impact of these variants on CAD, we then performed logistic regression analysis, adjusted for age at recruitment and sex, on the CAD status between noncarriers and carriers of all strong *ANGPTL3* PTVs and obtained odds ratios for the effect of the PTVs on the risk of prevalent CAD by exponentiating the logistic regression coefficient. In order to obtain a positive control for this natural experiment, the same analytical strategy was performed for PTVs in *PCSK9*, except that variants were considered strong if they were significantly associated with a reduction in the levels of circulating apoB instead of triglycerides.

### Data and code availability

R version 4.0.4 and RStudio Server 1.3.1093 were used to analyze data and create plots^50^. TwoSampleMR 0.5.6 (https://github.com/MRCIEU/TwoSampleMR)^51^, the MendelianRandomization 0.5.1 (https://cran.r-project.org/web/packages/ MendelianRandomization), the ieugwasr (https://mrcieu.github.io/ieugwasr/), the data.table 1.14.0 (https://github. com/Rdatatable/data.table), the coloc (https://cran.r-project.org/ web/packages/coloc/) and the HyPrColoc (https://github.com/ jrs95/hyprcoloc) packages in R. The alignment pipeline is available at https://github.com/broadinstitute/gtex-pipeline/tree/master/rnaseq. Code for analysis is available at the associated website of each software package. All genome-wide association study summary statistics used are publicly available. Summary statistics for ANGPTL3 levels from deCODE genetics were downloaded from https://www.decode.com/summarydata/. The full summary statistics for the genome-proteome-wide association study for ANGPTL3 from the Fenland cohort were downloaded from: https://omicscience<u>. org/apps/pgwas/</u>.

**Table 1.**
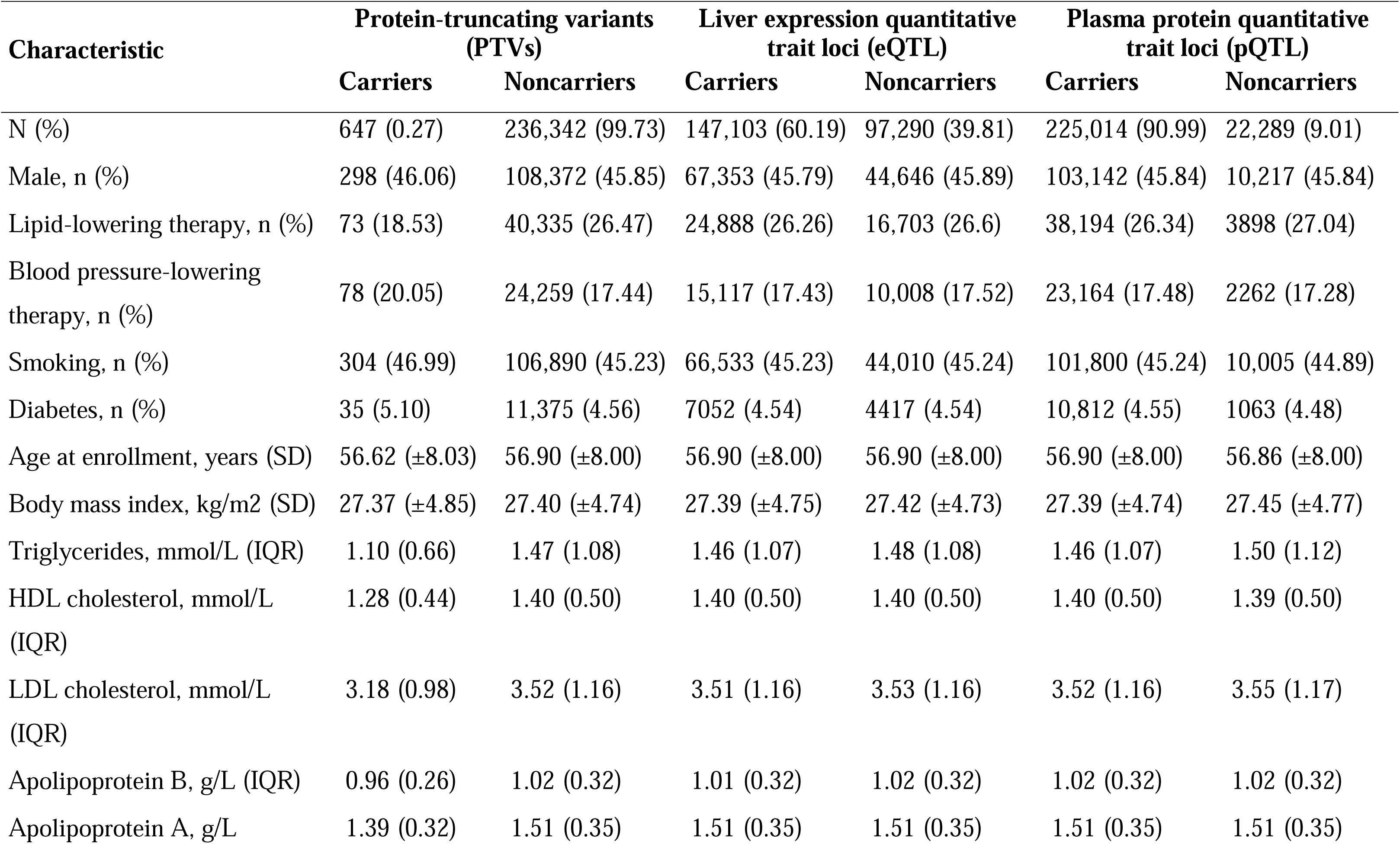
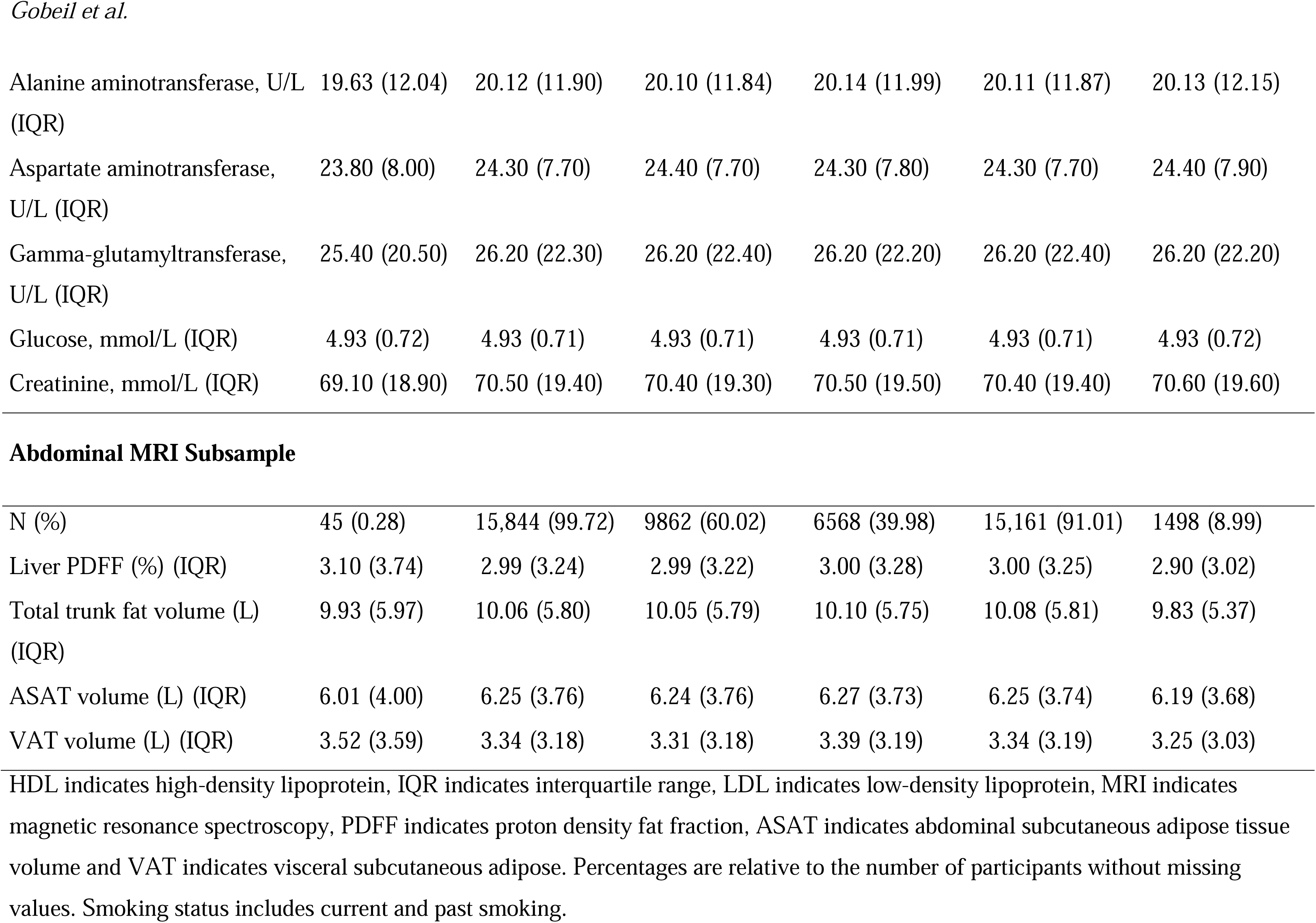
Baseline characteristics of the UK Biobank participants according to carrier status of *ANGPTL3* variants linked with lower plasma triglyceride levels.

| Characteristic | Protein-truncating variants (PTVs) |  | Liver expression quantitative trait loci (eQTL) |  | Plasma protein quantitative trait loci (pQTL) |  |
| --- | --- | --- | --- | --- | --- | --- |
|  | Carriers | Noncarriers | Carriers | Noncarriers | Carriers | Noncarriers |
| N (%) | 647 (0.27) | 236,342 (99.73) | 147,103 (60.19) | 97,290 (39.81) | 225,014 (90.99) | 22,289 (9.01) |
| Male, n (%) | 298 (46.06) | 108,372 (45.85) | 67,353 (45.79) | 44,646 (45.89) | 103,142 (45.84) | 10,217 (45.84) |
| Lipid-lowering therapy, n (%) | 73 (18.53) | 40,335 (26.47) | 24,888 (26.26) | 16,703 (26.6) | 38,194 (26.34) | 3898 (27.04) |
| Blood pressure-lowering therapy, n (%) | 78 (20.05) | 24,259 (17.44) | 15,117 (17.43) | 10,008 (17.52) | 23,164 (17.48) | 2262 (17.28) |
| Smoking, n (%) | 304 (46.99) | 106,890 (45.23) | 66,533 (45.23) | 44,010 (45.24) | 101,800 (45.24) | 10,005 (44.89) |
| Diabetes, n (%) | 35 (5.10) | 11,375 (4.56) | 7052 (4.54) | 4417 (4.54) | 10,812 (4.55) | 1063 (4.48) |
| Age at enrollment, years (SD) | 56.62 ($\pm 8.03$) | 56.90 ($\pm 8.00$) | 56.90 ($\pm 8.00$) | 56.90 ($\pm 8.00$) | 56.90 ($\pm 8.00$) | 56.86 ($\pm 8.00$) |
| Body mass index, kg/m<sup>2</sup> (SD) | 27.37 ($\pm 4.85$) | 27.40 ($\pm 4.74$) | 27.39 ($\pm 4.75$) | 27.42 ($\pm 4.73$) | 27.39 ($\pm 4.74$) | 27.45 ($\pm 4.77$) |
| Triglycerides, mmol/L (IQR) | 1.10 (0.66) | 1.47 (1.08) | 1.46 (1.07) | 1.48 (1.08) | 1.46 (1.07) | 1.50 (1.12) |
| HDL cholesterol, mmol/L (IQR) | 1.28 (0.44) | 1.40 (0.50) | 1.40 (0.50) | 1.40 (0.50) | 1.40 (0.50) | 1.39 (0.50) |
| LDL cholesterol, mmol/L (IQR) | 3.18 (0.98) | 3.52 (1.16) | 3.51 (1.16) | 3.53 (1.16) | 3.52 (1.16) | 3.55 (1.17) |
| Apolipoprotein B, g/L (IQR) | 0.96 (0.26) | 1.02 (0.32) | 1.01 (0.32) | 1.02 (0.32) | 1.02 (0.32) | 1.02 (0.32) |
| Apolipoprotein A, g/L | 1.39 (0.32) | 1.51 (0.35) | 1.51 (0.35) | 1.51 (0.35) | 1.51 (0.35) | 1.51 (0.35) |
| Alanine aminotransferase, U/L (IQR) | 19.63 (12.04) | 20.12 (11.90) | 20.10 (11.84) | 20.14 (11.99) | 20.11 (11.87) | 20.13 (12.15) |
| Aspartate aminotransferase, U/L (IQR) | 23.80 (8.00) | 24.30 (7.70) | 24.40 (7.70) | 24.30 (7.80) | 24.30 (7.70) | 24.40 (7.90) |
| Gamma-glutamyltransferase, U/L (IQR) | 25.40 (20.50) | 26.20 (22.30) | 26.20 (22.40) | 26.20 (22.20) | 26.20 (22.40) | 26.20 (22.20) |
| Glucose, mmol/L (IQR) | 4.93 (0.72) | 4.93 (0.71) | 4.93 (0.71) | 4.93 (0.71) | 4.93 (0.71) | 4.93 (0.72) |
| Creatinine, mmol/L (IQR) | 69.10 (18.90) | 70.50 (19.40) | 70.40 (19.30) | 70.50 (19.50) | 70.40 (19.40) | 70.60 (19.60) |
| Abdominal MRI Subsample |  |  |  |  |  |  |
| N (%) | 45 (0.28) | 15,844 (99.72) | 9862 (60.02) | 6568 (39.98) | 15,161 (91.01) | 1498 (8.99) |
| Liver PDFF (%) (IQR) | 3.10 (3.74) | 2.99 (3.24) | 2.99 (3.22) | 3.00 (3.28) | 3.00 (3.25) | 2.90 (3.02) |
| Total trunk fat volume (L) (IQR) | 9.93 (5.97) | 10.06 (5.80) | 10.05 (5.79) | 10.10 (5.75) | 10.08 (5.81) | 9.83 (5.37) |
| ASAT volume (L) (IQR) | 6.01 (4.00) | 6.25 (3.76) | 6.24 (3.76) | 6.27 (3.73) | 6.25 (3.74) | 6.19 (3.68) |
| VAT volume (L) (IQR) | 3.52 (3.59) | 3.34 (3.18) | 3.31 (3.18) | 3.39 (3.19) | 3.34 (3.19) | 3.25 (3.03) |

HDL indicates high-density lipoprotein, IQR indicates interquartile range, LDL indicates low-density lipoprotein, MRI indicates magnetic resonance spectroscopy, PDFF indicates proton density fat fraction, ASAT indicates abdominal subcutaneous adipose tissue volume and VAT indicates visceral subcutaneous adipose. Percentages are relative to the number of participants without missing values. Smoking status includes current and past smoking.

## Results

The schematic representation of the study design is presented in Figure 1. The overarching objective of this investigation was to estimate the health effects of multiple ANGPTL3 inhibition strategies by mimicking the impact of these strategies using naturally occurring genetic variations. We mimicked the effect of a liver-targeted, RNA-based *ANGPTL3* inhibition strategy by determining the cardiometabolic effects of a variant associated with lifelong exposure to lower liver *ANGPTL3* gene expression levels (a liver expression quantitative trait locus). We mimicked the effect of an antibody-based ANGPTL3 inhibition strategy in the blood by determining the cardiometabolic effects of a variant (or a combination of variants) associated with lifelong exposure to lower plasma ANGPTL3 protein levels. We also mimicked the effect of gene editing-based ANGPTL3 inhibition strategy by determining the cardiometabolic effects of sequence variants associated with lifelong exposure to lower plasma triglyceride levels due to a truncated ANGPTL3 protein.

**Figure 1.**
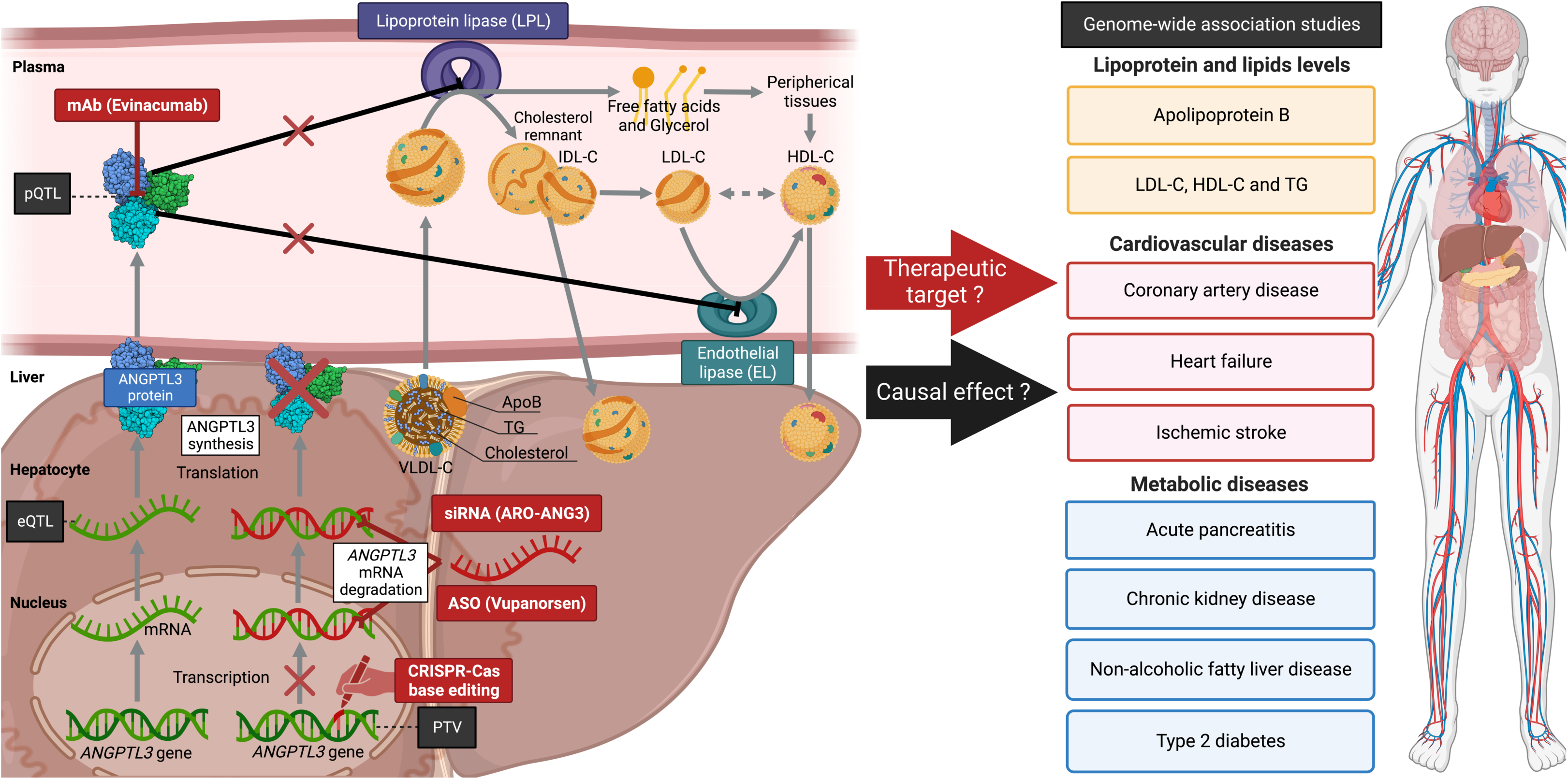
Schematic representation of the Mendelian randomization study design. ANGPTL3 is a key regulator of blood lipid levels. ANGPTL3 inhibitors are currently under investigation for their impact on metabolic and cardiovascular health. ANGPTL3: angiopoietin-like protein-3; ApoB: apolipoprotein B; ASO: antisense oligonucleotides; eQTL: expression quantitative trait loci; HDL-C: high-density lipoprotein cholesterol; IDL-C: intermediate-density lipoprotein cholesterol; LDL-C: low-density lipoprotein cholesterol; mAb: monoclonal antibody; mRNA: messenger ribonucleic acid; pQTL: protein quantitative trait loci; PTV: protein-truncating variants; siRNA: small interfering RNA; TG: triglycerides; VLDL-C: very-low-density lipoprotein cholesterol.

### Liver ANGPTL3 expression and cardiometabolic traits and diseases

In order to determine whether genetic variants influence liver expression levels of *ANGPTL3*, we performed RNA sequencing of liver samples obtained from 246 patients undergoing bariatric surgery (IUCPQ Obesity Biobank). One variant in the *ANGPTL3* locus (rs11207978) had a strong effect on the liver *ANGPTL3* expression (p=25×10^-6^). We investigated the impact of this variant on lipoprotein-lipid levels and cardiometabolic outcomes using MR and genetic colocalization. As presented in Figure 2 and Supplementary Table 4, this SNP has a very strong effect on plasma triglyceride levels, a more modest effect on LDL cholesterol and a weaker effect of apoB levels. However, this variant was not associated with CAD and other cardiometabolic diseases.

**Figure 2.**
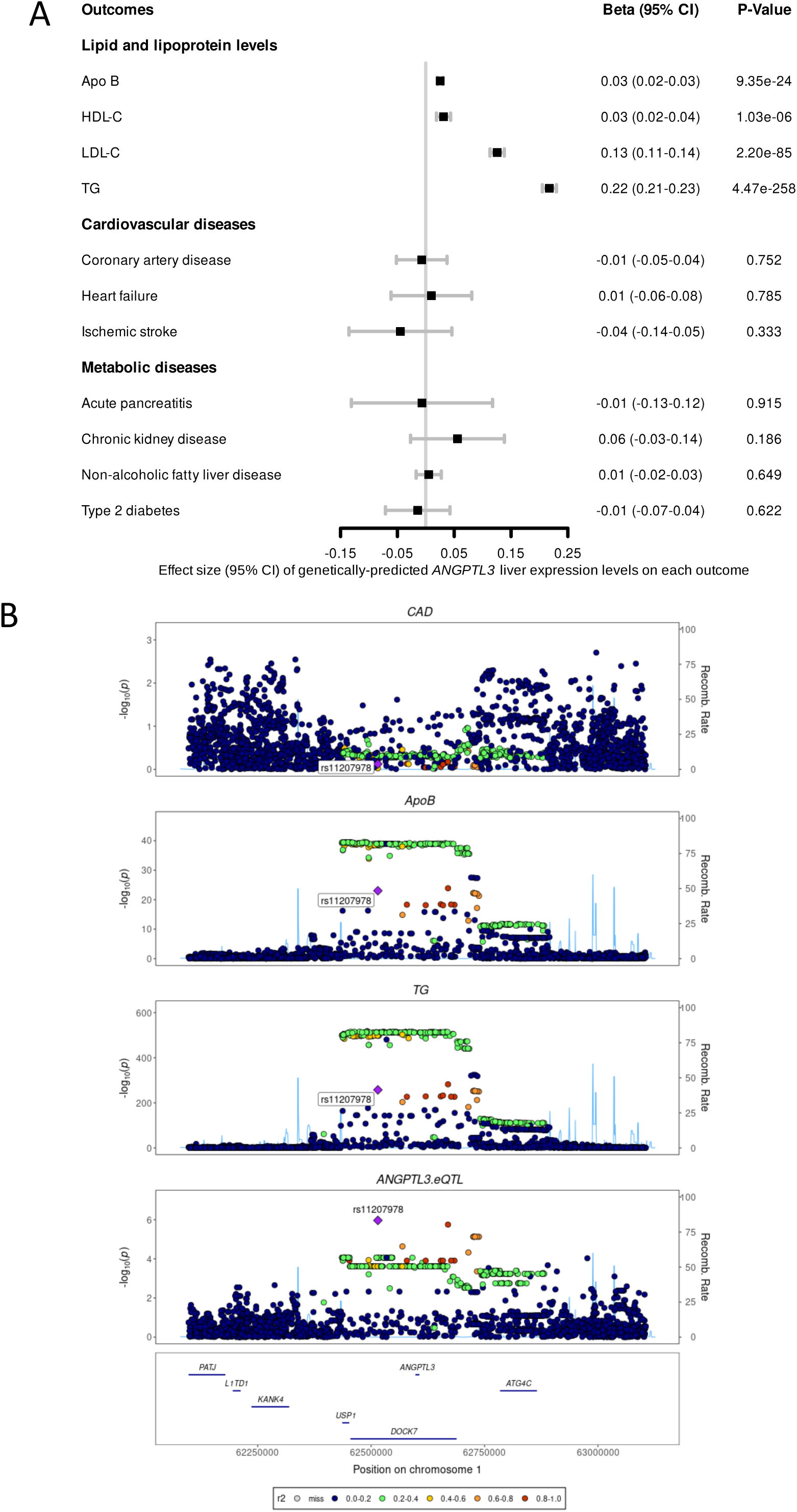
Impact of genetically-predicted liver *ANGPTL3* expression on lipoprotein-lipid levels and cardiometabolic diseases. A) The effects of the strongest SNP (rs11207978) associated with the liver *ANGPTL3* expression on lipoprotein-lipid levels and cardiometabolic diseases was obtained using the Wald ratio. Effect sizes (95% CI) are represented by SD or log(OR) change in the outcome per 1-SD increase in liver *ANGPTL3* expression. B) Regional association plots depicting the impact of genetically-predicted liver *ANGPTL3* expression on lipoprotein-lipid levels and coronary artery disease.

### ANGPTL3 blood levels and cardiometabolic traits and diseases

To determine whether genetic variants influencing blood levels of ANGPTL3, we investigated the impact of the top variant in the *ANGPTL3* locus influencing blood levels of ANGPTL3 in the deCODE cohort (rs10889352) on lipoprotein-lipid levels and cardiometabolic outcomes using MR and genetic colocalization. Figure 3 and Supplementary Table 5 show the association between the top ANGPTL3 blood pQTL and lipoprotein-lipid levels and cardiometabolic diseases. This variant has a very strong effect on plasma triglyceride levels, a more modest effect on LDL cholesterol and a weaker effect of apoB levels. This variant was not associated with CAD and other cardiometabolic diseases.

**Figure 3.**
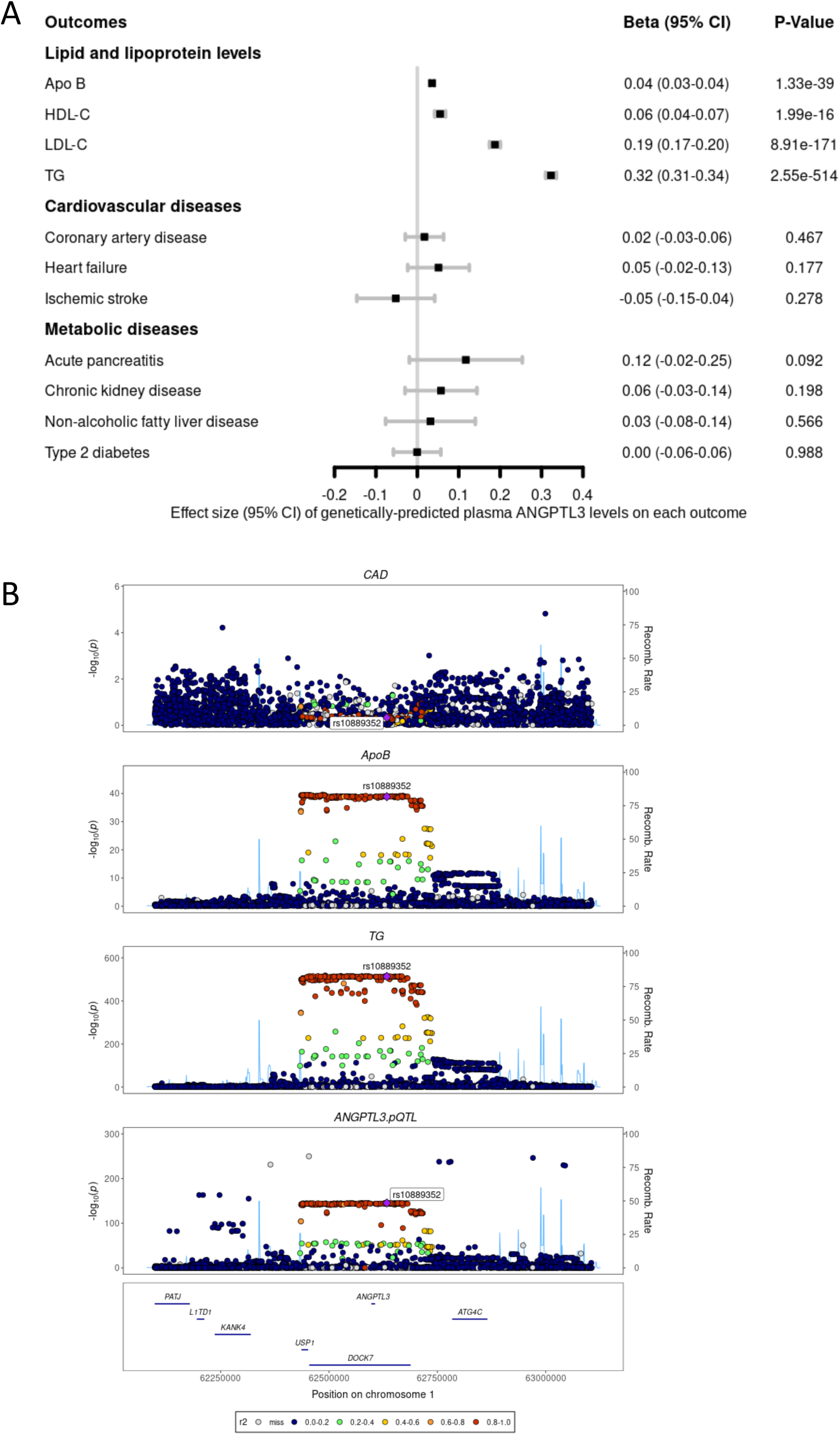
Impact of genetically-predicted plasma ANGPTL3 levels on lipoprotein-lipid levels and cardiometabolic diseases. **A)** The effects of the strongest SNP (rs10889352) associated with the plasma ANGPTL3 levels on lipoprotein-lipid levels and cardiometabolic diseases was obtained using the Wald ratio. Effect sizes (95% CI) are represented by SD or log(OR) change in the outcome per 1-SD increase in plasma *ANGPTL3* levels. B) Regional association plots depicting the impact of genetically-predicted plasma *ANGPTL3* levels on lipoprotein-lipid levels and coronary artery disease.

### Genome-wide association studies of circulating ANGPTL3 levels

To identify and validate genetic loci that may influence circulating levels of ANGPTL3 and that could be used in a more comprehensive MR setting, we investigated the results of the deCODE and Fenland studies, which measured ANGPTL3 levels in 35,359 and 10,078 individuals, respectively. Figure 4 presents the results of the GWAS for ANGPTL3 levels in these two cohorts. In deCODE, genetic variation at five loci including *ANGPTL3*, *LPL* (the gene encoding lipoprotein lipase), *APOA5-APOA4-APOC3-APOA1*, *ANGPTL4* and *APOE* were found to influence ANGPTL3 levels at the genome-wide significant level (p<5e-08). The Fenland cohort replicated three of these loci at genome-wide level and two of these loci at p<0.01.

**Figure 4.**
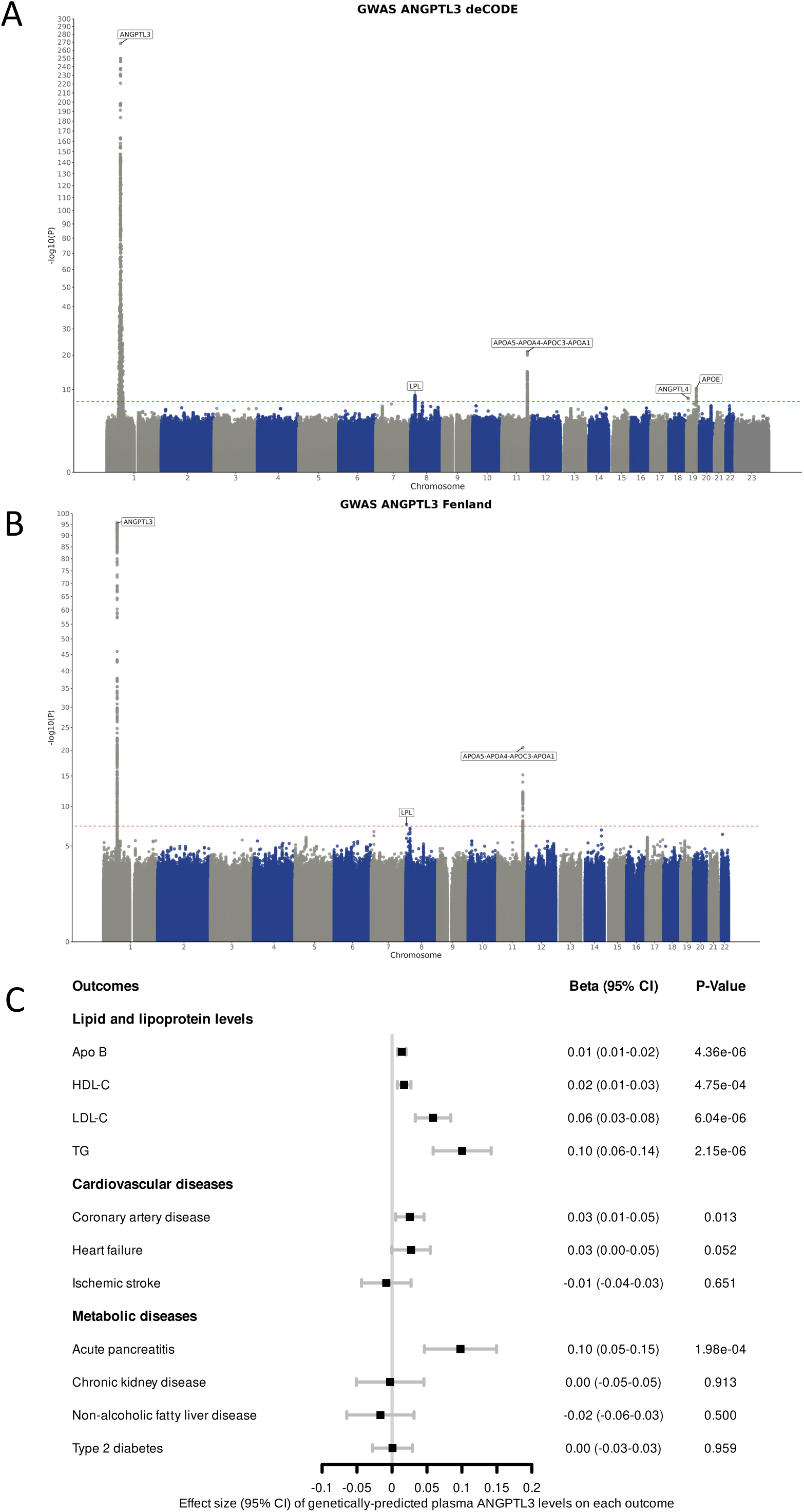
Genome-wide association studies of circulating angiopoietin-like protein-3 levels. Manhattan plot representing the results of the genome-wide association studies of circulating angiopoietin-like protein-3 levels in the A) deCODE and B) Fenland cohorts. C) The effects of single-nucleotide polymorphisms associated with circulating ANGPTL3 levels on lipoprotein-lipid levels and cardiometabolic diseases was obtained using inverse-variance weighted Mendelian randomization. Effect sizes (95% CI) are represented by SD or log(OR) change in the outcome per 1-SD increase in circulating ANGPTL3 levels.

Supplementary Table 6 presents the effect size and characteristics of the top variant influencing ANGPTL3 levels at these loci in both cohorts. To investigate tissue specificity of the genes influencing ANGPTL3 levels, we assessed genetic cololalization using visceral and subcutaneous adipose tissue samples from the GTEx project as well as liver samples from the IUCPQ Obesity Biobank. The signal at the *ANGPTL3* and *APOE* loci colocalized with liver expression of *ANGPTL3* and the signal at the *LPL* locus colocalized with subcutaneous expression of *LPL* (Supplementary Table 7). Results of this analysis suggest that genes involved in triglyceride-rich lipoprotein metabolism that are expressed in the liver or subcutaneous adipose tissue influence circulating concentrations of ANGPTL3. Variants influencing plasma ANGPTL3 levels were all involved in triglyceride-rich lipoprotein metabolism. To avoid horizontal pleiotropy, we have opted to only include SNPs in the *ANGPTL3* locus for further MR analyses. Using independent genetic instruments (Supplementary Table 3), we further investigated the impact of genetically-predicted ANGPTL3 levels with the inverse variance-weighted MR method. These results show that blood ANGPLT3 levels have a strong effect on plasma triglyceride levels, a more modest effect on LDL cholesterol and a weaker effect of apoB levels (Figure 4C). Genetically-predicted ANGPTL3 levels were associated with acute pancreatitis. Genetically-predicted ANGPTL3 blood levels were also associated with coronary artery disease but the association was weak and not significant after adjusting for multiple testing (p<0.05/11=0.0045) (Supplementary Table 8).

### Genetically-predicted triglyceride levels and apolipoprotein B levels

Results of several recent MR studies and clinical trials provided evidence that lipid-lowering therapies may be effective in reducing ASCVD risk if they provide significant reductions in plasma apo B levels (regardless of their impact on other lipoprotein parameters such as triglyceride or HDL cholesterol levels). Given that variants influencing liver expression of *ANGPTL3* or blood protein levels of ANGPTL3 had a strong effect on triglyceride levels but a weaker effect on apoB, we investigated if the reductions in triglycerides predicted reductions in apoB and if these reductions were differently predicted if they were produced by changes in ANGPTL3 levels. Figure 5A presents the result of a MR study on the effects of triglyceride levels on apolipoprotein B levels with variants in the *ANGPTL3* locus presented in red. Results of this analysis suggest a causal effect of triglycerides on apoB levels. Since variants in the *ANGPTL3* region are either on the regression line or very close to it, it may be concluded that the effect of reductions in triglycerides can predict reductions in apoB levels and that reductions in triglycerides following inhibition of ANGPTL3 levels predict reductions in apoB levels as would be expected by most triglyceride-lowering mechanisms.

**Figure 5.**
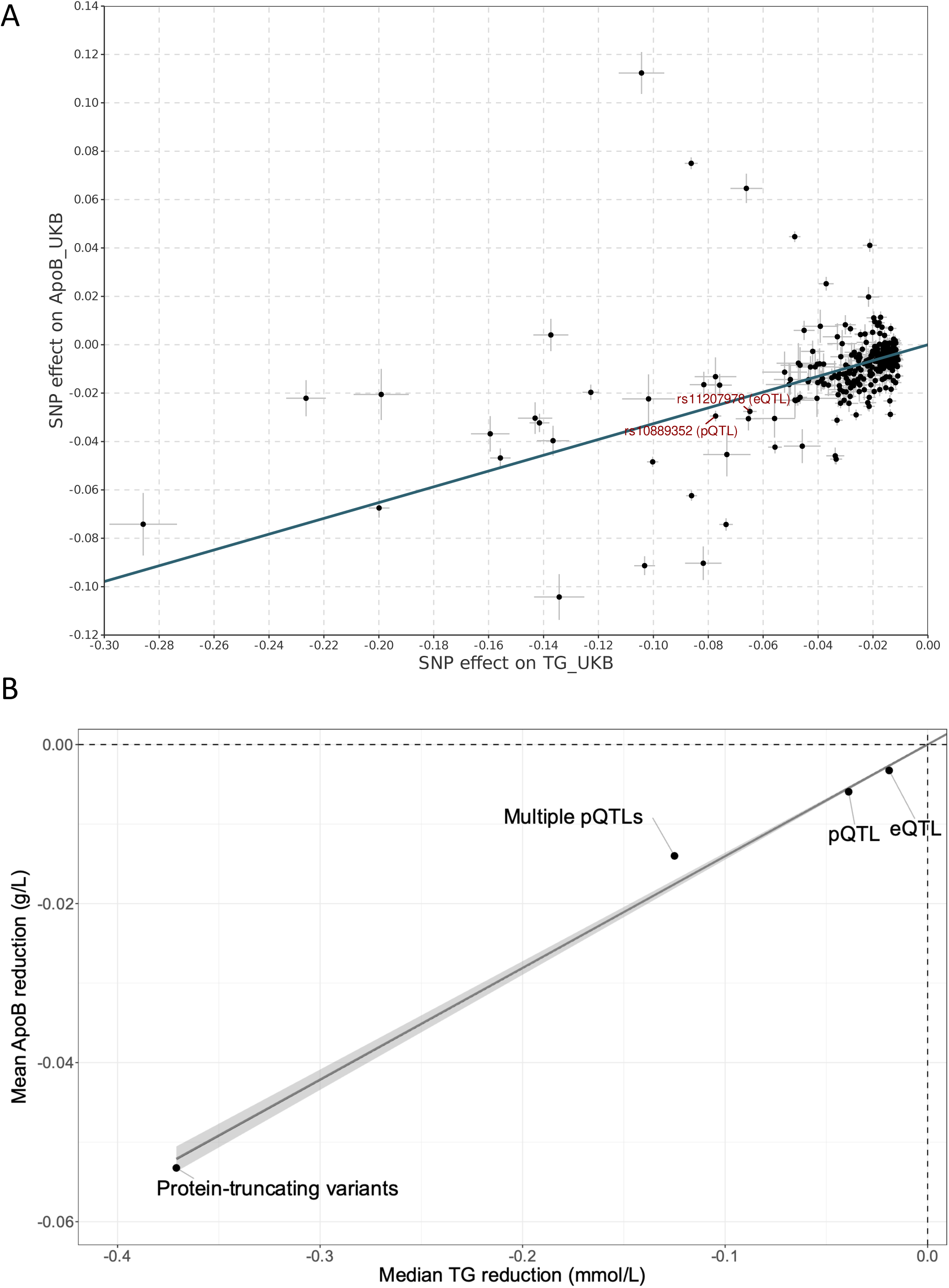
Impact of genetic variation at *ANGPTL3* on triglyceride and apolipoprotein B levels. A) Inverse-variance-weighted Mendelian randomization scatterplot depicting the causal impact of triglyceride-reducing SNPs on apolipoprotein B (apoB) levels. The top liver eQTL-SNP (rs11207978) influencing *ANGPTL3* hepatic gene expression levels and the top plasma pQTL-SNP (rs10889352) influencing ANGPTL3 levels are highlighted. B) Impact of rare and common genetic variants at the *ANGPTL3* locus on triglyceride and apoB reductions.

### Impact of ANGPTL3 protein-truncating variants on triglyceride levels

Using whole-exome sequencing data on 407,668 UK Biobank participants, we identified 647 participants carrying at least one of four ANGPTL3 PTVs associated with lower plasma triglyceride levels (Table 1). These PTVs have a stronger effect on both triglyceride and apoB levels compared to the top eQTLs, the top pQTL and combination of pQTLs. Carriers of PTVs also had a comparable cardiometabolic risk profile including similar body fat distribution patterns as noncarriers. As presented in Figure 5B, the top liver eQTL-SNP (rs11207978) influencing *ANGPTL3* hepatic gene expression levels, the top plasma pQTL-SNP (rs10889352) influencing ANGPTL3 levels, the combination of independent pQTLs influencing ANGPTL3 levels (per 1-SD decrease in ANGPTL3 levels) as well as *ANGPTL3* PTV provide apoB reductions that can be predicted by their reduction in triglyceride levels. Despite having a notable effect on triglyceride levels (-0.37 [interquartile range=0.41] mmol/L), these variants showed a modest effect on apoB levels (-0.06±0.32] g/L). Characteristics and individuals’ estimates of ANGPTL3 PTVs associated with lower triglyceride levels in participants of the UK Biobank are presented in Supplementary Table 9 and Supplementary Table 10. Altogether, these results suggest that genetic variants at the *ANGPTL3* locus are associated with small reductions in apoB levels and that the reductions in apoB are proportional to the reduction in triglyceride levels.

### Impact of ANGPTL3 protein-truncating variants on coronary artery disease

We finally sought to investigate whether participants of the UK Biobank carrying at least one ANGPTL3 PTVs associated with plasma triglyceride levels had a lower rate of CAD compared to noncarriers. In this population, the CAD event rate was comparable in carriers versus noncarriers (Figure 6). To present a positive control for this “natural” experiment, we investigated whether carriers with at least one PCSK9 PTVs associated with plasma apoB levels (N=475) had a lower rate of CAD compared to noncarriers. As expected, carriers of sequence variants at the PCSK9 locus associated with lower apoB levels had a lower CAD event rate compared to noncarriers. Altogether, results of this analysis suggest that despite having important lifelong reduction in triglyceride levels, carriers of *ANGPTL3* PTVs have smaller lifelong reductions in apoB and a similar CAD risk compared to noncarriers.

**Figure 6.**
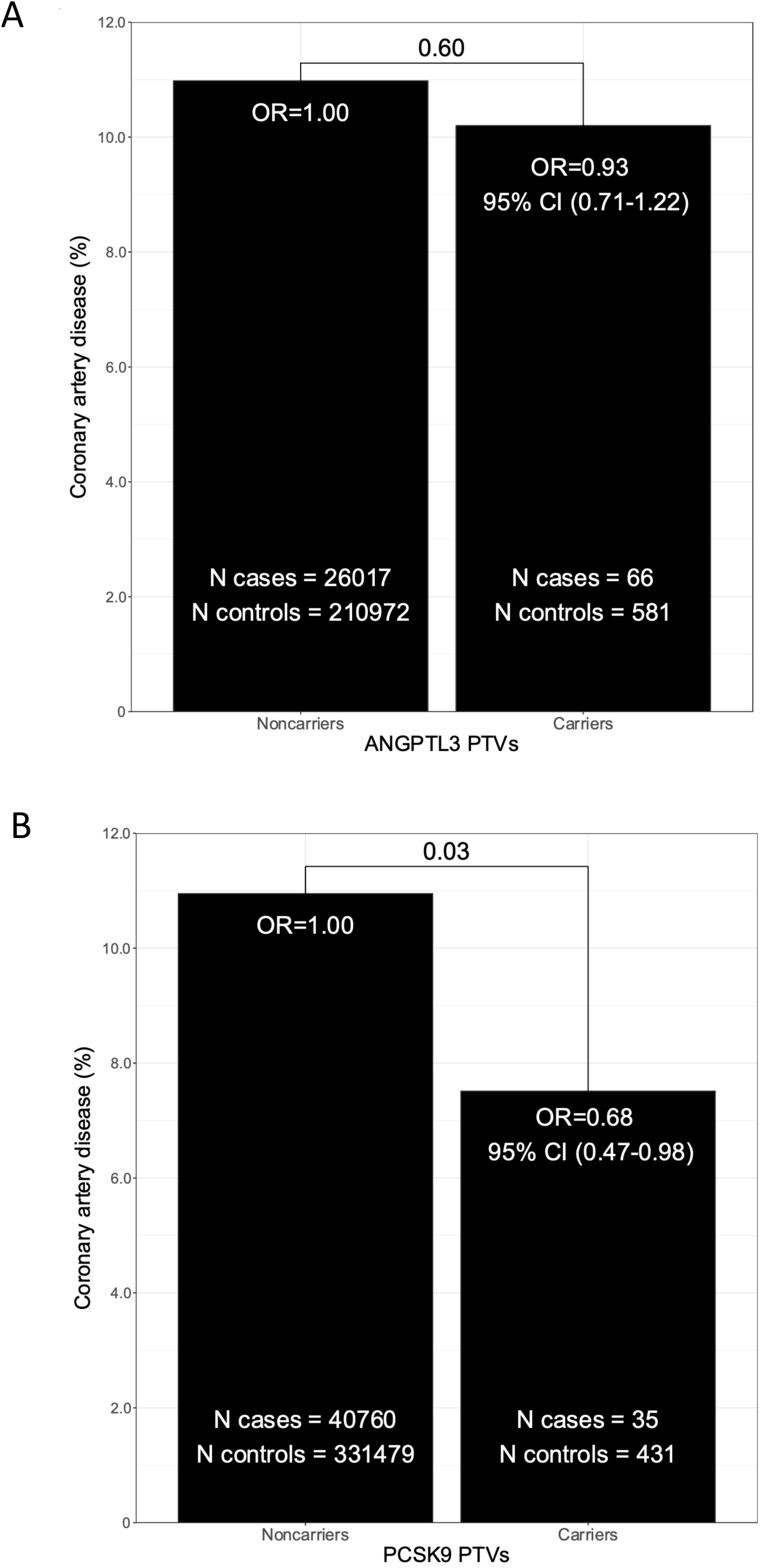
Impact of *ANGPTL3* protein-truncating variants on coronary artery disease rates in participants of the UK Biobank. A) Carriers of at least one *ANGPTL3* protein-truncating variant (PTV) associated with lower plasma triglyceride levels had a comparable coronary artery disease (CAD) event rate than noncarriers. B) Carriers of at least one *PCSK9* PTV associated with lower plasma apolipoprotein B levels had a lower CAD event rate than noncarriers.

## Discussion

In this study, we leveraged multiple genetic strategies to explore the potential health effects of reducing triglyceride levels through ANGPTL3 inhibition. Using multiple MR strategies, we mimicked the effect of a liver-targeted, RNA-based *ANGPTL3* inhibition strategy, the effect of an antibody-based ANGPTL3 inhibition strategy as well as the effect of gene editing-based *ANGPTL3* inhibition strategy. These analyses provided consistent evidence that ANGPTL3 “genetic inhibition” had a strong effect on circulating triglyceride levels, a weak effect on circulating apoB levels and no effect on CAD. Although the impact of ANGPTL3-mediated triglyceride reductions on ASCVD will ultimately need to be examined in large cardiovascular outcomes trials, our findings suggest that the long-term cardiometabolic benefits of this therapeutic strategy in patients at high residual risk of ASCVD with elevated triglyceride levels may be limited.

Cardiovascular outcome trials and MR studies have provided convincing evidence that lipid-lowering therapies may reduce long-term ASCVD risk if they provide significant reductions in plasma apoB levels and that the reduction in apoB levels is proportional to ASCVD risk reductions^5, 6, 52^. Our MR findings suggest that although ANGPTL3 inhibition may profoundly reduce plasma triglyceride levels, this strategy provides small apoB reductions. This finding is consistent with the results of randomized clinical trials^14, 15, 53^. In patients with elevated triglyceride levels, T2D and NAFLD, treatment with the liver-targeted antisense oligonucleotide against *ANGPTL3* vupanorsen administered once every four week for six months provided large reductions in triglyceride levels (53%) and small reductions in apoB (9%) at the dose of 80 mg^14^. In the TRANSLATE-TIMI 70 trial, a higher exposure to vupanorsen (160 mg administered once every four week for 24 weeks) provided large triglyceride reductions (45.9%) and also smaller apoB reductions (12.6%) in statin-treated patients with elevated triglyceride and non-HDL cholesterol levels^15^. In smaller phase 1 trials performed in patients with elevated triglycerides and LDL cholesterol, treatment with the monoclonal antibody evinacumab provided more potent short-term effects^53^. In a single escalating dose study, maximal triglyceride reduction (76.9%) was observed on day 3 with intravenous evinacumab (10 mg/kg) while the maximal triglyceride reduction (83.1%) was observed on day 2 in a multiple escalating dose study. While the corresponding reductions in apoB at these timepoints were approximately 10% and 20%, respectively, more important apoB reductions (reaching a nadir of 25% and 35%, respectively) were observed in the following days. Subcutaneous evinacumab injections provided consistent but less potent effects. Altogether, our study, combined with those of randomized clinical trials of short-term ANGPTL3 inhibition with RNA-based and antibody-based therapies suggest that very potent and near-complete ANGPTL3 reductions will likely be required to reducing significantly apoB levels and ultimately ASCVD risk. Since we also observed that variants influencing triglyceride levels in the *ANGPTL3* region predict apoB reductions similarly to the predicted impact of an average triglyceride gene, it may be concluded that reductions in triglycerides following inhibition of ANGPTL3 would predict reductions in apoB levels as would be expected by most triglyceride-lowering mechanisms. Exploring other triglyceride-reduction mechanisms that provide more potent apoB reductions than what could be expected by reductions in triglyceride levels need to be explored to provide ASCVD-risk reductions in patients with high triglycerides with an elevated ASCVD residual risk.

Although our study provides evidence that ANGPTL3-mediated triglyceride reductions may have a limited impact on apoB and ASCVD, this study does not rule out potential benefits of ANGPTL3 inhibition in other patients populations such as familial hypercholesterolemia (FH). In patients with heterozygous FH with refractory hypercholesterolemia or homozygous FH, treatment with evinacumab reduced LDL cholesterol levels by approximately 50% and provided apoB reductions in the same range^16, 54^. Interestingly, individuals carrying a loss-of-function mutation in the *ANGPTL3* gene and low LDL cholesterol levels do have a lower CAD risk compared to noncarriers^24^. In our study, only one PTV was found to be associated with apoB levels and the association of this PTV with CAD could not be explored since this PTV was found in only six individuals. Our study does not rule out the possibility that ANGPTL3 inhibitors, regardless of inhibition strategy, could lead to important reductions in apoB levels and a reduction in ASCVD risk in patients with elevated LDL/apoB levels on optimal lipid-lowering therapy.

Interestingly, we found an effect of circulating ANGPTL3-mediated reductions in triglyceride levels on acute pancreatitis, suggesting that antibody-based reductions in triglycerides could prevent acute pancreatitis, a disease that is highly prevalent in patients with hypertriglyceridemia^55^. However, we could note explore whether ANGPTL3 PTVs were associated with acute pancreatitis because of low statistical power. We also found no associations between triglyceride-reducing *ANGPTL3* variants (by any mechanism) and liver fat accumulation, liver enzymes and NAFLD. This finding contrasts those of RNA-based therapies against ANGPTL3 that have reported increases in hepatic fat fractions in some^15^, but not all studies^14^. Whether increases in liver fat fraction following ANGPTL3 inhibition reflect a true biological phenomenon or an off-target effect of vupanorsen will need to be further explored in longer-term randomized clinical trials.

Results of this study also provide new information on the genetic and biological determinants of circulating ANGPTL3 levels. Through a combination of GWAS and genetic colocalization with our liver gene expression dataset as well as that of the Genotype-Tissue Expression (GTEx), we found that several genes outside of the *ANGPTL3* locus implicated in triglyceride-rich lipoprotein metabolism could influence circulating ANGPTL3 levels. For instance, signals colocalizing respectively with hepatic expression of *APOE* and subcutaneous adipose tissue expression of *LPL* were found. Other signals at the *ANGPTL4* as well as *APOA5-APOA4-APOC3-APOA1* loci were identified. This finding may suggest a bidirectional relationship between ANGPTL3 and triglyceride-rich metabolism. Indeed, although ANGPTL3 genetic or pharmacologic modulation does influence lipoprotein metabolism, other determinants of triglyceride-rich metabolism could in turn influence plasma ANGPTL3 levels. Whether these genes influence free or lipoprotein-bound ANGPTL3 needs to be further explored. Because these variants all involved in triglyceride-rich lipoprotein metabolism, they could introduce horizontal pleiotropy in MR analyses. Consequently, we only include SNPs in the *ANGPTL3* locus in our MR analyses aiming to predict long-term effects inhibition of ANGPTL3 with antibody-based strategies.

In conclusion, our study is the first to use a comprehensive MR study design to predict the long-term health effects of three modes of action evaluating the possible health benefits of a therapeutic target under investigation. Our results suggest that RNA-interfering-based, antibody-based, and gene-editing based ANGPTL3 inhibition provide important effects on circulating triglyceride levels, less potent effects on circulating apoB levels and no effects on CAD. Therefore, ANGPTL3-mediated triglyceride reductions may have a limited impact on apoB and ASCVD risk in patients with high triglycerides at high ASCVD risk despite optimal LDL/apoB levels. Near-complete inhibition of ANGPTL3 function, however, could lower apoB levels and potentially reduce ASCVD risk in patients with lipid disorders characterized by elevated concentrations of plasma apoB who are unable to meet guideline-recommended lipoprotein-lipid levels.

## Data Availability

R version 4.0.4 and RStudio Server 1.3.1093 was used to analyze data and create plots50. TwoSampleMR 0.5.6 (https://github.com/MRCIEU/TwoSampleMR)51, the MendelianRandomization 0.5.1 (https://cran.r-project.org/web/packages/MendelianRandomization), the ieugwasr (https://mrcieu.github.io/ieugwasr/), the data.table 1.14.0 (https://github.com/Rdatatable/data.table), the coloc (https://cran.r-project.org/web/packages/coloc/) and the HyPrColoc (https://github.com/jrs95/hyprcoloc) packages in R. The alignment pipeline is available at https://github.com/broadinstitute/gtex-pipeline/tree/master/rnaseq. Code for analysis is available at the associated website of each software package. All genome-wide association study summary statistics used are publicly available. Summary statistics for ANGPTL3 levels from deCODE genetics were downloaded from https://www.decode.com/summarydata/. The full summary statistics for the genome-proteome-wide association study for ANGPTL3 from the Fenland cohort were downloaded from: https://omicscience.org/apps/pgwas/.

## Acknowledgements

The authors would like to thank all study participants as well as all investigators of the studies that were used throughout the course of this investigation. We acknowledge the invaluable collaboration of the surgery team, bariatric surgeons, and biobank staff and would like to thank all study participants from the Institut universitaire de cardiologie et de pneumologie de Québec (IUCPQ).

## Funding

This study was funded by Pfizer with additional support from the Canadian Institutes of Health Research (CIHR) and the Foundation of the IUCPQ. The funders had no role in the design of the study; in the collection, analyses, or interpretation of data; in the writing of the manuscript, or in the decision to publish results. The Genotype-Tissue Expression (GTEx) Project was supported by the Common Fund of the Office of the Director of the National Institutes of Health, and by National Cancer Institute, National Human Genome Research Institute, National Heart, Lung and Blood Institute, National Institute on Drug Abuse, National Institute of Mental Health, and National Institute of Neurological Disorders and Stroke. ÉG hold a Doctoral Research Award from the CIHR. JB holds a Masters’ Research Award from the *Fonds de recherche du Québec: Santé* (FRQS). EG holds a Doctoral Research Award from the *FRQS*. BJA holds a Senior Scholar Award from the FRQS. YB holds a Canada Research Chair in Genomics of Heart and Lung Diseases. PM holds a FRQS Research Chair on the Pathobiology of Calcific Aortic Valve Disease. MCV is Canada Research Chair in Genomics applied to Nutrition and Metabolic Health. These authors disclose the following: BJA is a consultant for Novartis, Editas Medicine, and Silence Therapeutics and has received research contracts from Pfizer, Ionis Pharmaceuticals, and Silence Therapeutics. AT receives research funding from Johnson & Johnson, Medtronic, and G.I. windows for studies related to bariatric surgery as well as consulting fees from Bausch Health and Novo Nordisk. The remaining authors disclose no conflicts.

## Supplementary Tables

**ST1.** Clinical characteristics of participants of the IUCPQ Obesity Biobank cohort.

**ST2.** Description of the GWAS datasets used.

**ST3.** Instrumental variables of SNPs associated with circulating ANGPTL3 levels for multi-SNP MR analysis.

**ST4.** Mendelian randomization with a single cis-acting SNP of the impact of genetically-predicted liver *ANGPTL3* expression on lipoprotein-lipid levels and cardiometabolic diseases.

**ST5.** Mendelian randomization with a single cis-acting SNP of the impact of genetically-predicted plasma ANGPTL3 levels on lipoprotein-lipid levels and cardiometabolic diseases.

**ST6.** Associations of previously identified variants linked with ANGPTL3 levels in two published GWAS (deCODE genetics and Fenland cohorts).

**ST7.** Genetic colocalization and tissue-specific effects of the top genetic variants influencing circulating ANGPTL3 levels.

**ST8.** IVW-MR results for SNPs associated with circulating ANGPTL3 levels on lipoprotein-lipid levels and cardiometabolic diseases.

**ST9.** Characteristics of *ANGPTL3* protein-truncating variants associated with lower triglycerides levels in participants of the UK Biobank.

**ST10.** Linear regression estimates of *ANGPTL3* protein-truncating variants associated with lower triglycerides levels in participants of the UK Biobank.

## Notes

### Author Declarations

The Ethics committee/IRB of Institut Universitaire de Cardiologie et de Pneumologie de Québec -Université Laval gave ethical approval for this work (Number of approval: 2021-3656, 22070).

