## Supplementary Methods for "Genetic inhibition of angiopoietin-like protein-3, lipoprotein-lipid levels and cardiometabolic diseases"

**Additional Methods**

Benoit Arsenault, PhD

Centre de recherche de l’Institut universitaire de cardiologie et de pneumologie de Québec – Université Laval

Y-3106, Pavillon Marguerite D'Youville, 2725 chemin Ste-Foy, Québec (QC), Canada G1V 4G5

**Aditionnal Methods**

*Identification of genetic variants influencing liver ANGPTL3 expression*

**DNA extraction and genome-wide genotyping**. Genomic DNA was extracted from the blood buffy coat using the GenElute Blood Genomic DNA kit (Sigma, St. Louis, MO, USA). Genome-wide genotyping was performed using the GSA-MD array (Illumina, San Diego, CA, USA). Hyb Oven at 37 degrees for 20-24 hours for amplification. Fragmentation was performed in a SciGene heat block for 1h at 37 Degrees. Denaturation is done in a SciGene heat block for 20 min at 95 Degrees, the plate was placed at room temperature for 30 min prior to hybridization. A total of 15 µL of resuspended DNA was then loaded on the array (placed on hybridization tray) and incubated in Illumina Hyb Oven at 48 degrees for hybridization for 16-20 hours. Staining was done using a Tecan Freedom Evo with LCG program. The Illumina iScan instrument was used for chip scanning. Quality controls were done using Illumina Genomestudio software 2.0 with Genotyping module version 2.0.5, with the auto-clustering option. Quality controls were performed using PlinkQC. Pre-imputation QC was made with R package plinkQC v0.3.4 and Will Rayner tools. The steps for pre-imputation QC included sex checking based on the homozygosity rate (<0.2 for females and >0.8 for males, participants with missing genotype (subject removed if greater than 3%), heterozygosity rate (participant removed if more than 3 SD from the mean, related participants (one subject removed if IBD PI_HAT from a pair > 0.1875, participants of divergent ancestry based on 1000G reference panel (participant removed if Euclidean distance from the center for Europeans is greater than 2), variants with excessive missingness rate (variant removed if missingness greater than 3%), variants with strong deviation from HWE (variant removed if difference vs observed and expected heterozygote frequencies has p < 1e-05), variants with low minor allele frequency (MAF < 0.025) were removed). Rayner tools were used to perform strand alignment with reference panel. Variants not in panel were removed. When comparing the study with reference panels, variants were removed if alleles were different or allele frequency differences greater than 0.2. Palindromic variants (A/T and G/C) with MAF > 0.4 were removed. Imputation was performed using the Michigan Imputation Server based on HRC reference panel. Eagle v2.4 and Minimac4 v1.6.3 were used for phasing and imputation respectively. Post-imputation QC steps included the creation of binary plink files with PLINK2.

**RNA extraction**. Total RNA was extracted from 10 mg of frozen liver samples following the manufacturer’s instructions (Qiagen RNeasy Plus). Briefly, tissue was homogenized in 800 µL Qiazol using a VWR Beadmill. A genomic DNA cleaning step was carried out followed by extraction. The concentration and purity (A260/A280 and A260/A230) of the samples were determined on a BioTech BioDrop. The quality of the RNA was determined either by BioAnalyzer or estimated by RNA agarose gel.

**RNA sequencing**. Total RNA was quantified, and quality assessed using a LabChip GXII (PerkinElmer). Libraries were generated from 250 ng of total RNA as following: mRNA enrichment was performed using the NEBNext Poly(A) Magnetic Isolation Module (New England BioLabs). cDNA synthesis was achieved with the NEBNext RNA First Strand Synthesis and NEBNext Ultra Directional RNA Second Strand Synthesis Modules (New England BioLabs). The remaining steps of library preparation were done using and the NEBNext Ultra II DNA Library Prep Kit for Illumina (New England BioLabs). Adapters and PCR primers were purchased from New England BioLabs. Libraries were quantified using the Kapa Illumina GA with Revised Primers-SYBR Fast Universal kit (Kapa Biosystems). Average size fragment was determined using a LabChip GXII (PerkinElmer) instrument. The libraries were normalized and pooled and then denatured in 0.05N NaOH and neutralized using HT1 buffer. The pool was loaded at 225pM on Illumina NovaSeq S4 lane using Xp protocol as per the manufacturer’s recommendations. The run was performed for 2x100 cycles (paired-end mode). A phiX library was used as a control and mixed with libraries at 1% level. Base calling was performed with RTA v3.4.4. Program bcl2fastq2 v2.20 was then used to demultiplex samples and generate fastq reads.

**RNA sequencing alignment**. Alignment to the human reference genome GRCh38/hg38 was performed using STAR v2.5.1, based on the GENCODE v34 annotation. The unaligned reads were kept in the final BAM file. Among multi-mapping reads, one read is flagged as the primary alignment by STAR.

**Transcript expression quantification**. Gene-level expression quantification was based on the GENCODE 34 annotation, collapsed to a single transcript model for each gene using a custom isoform collapsing procedure, including the following steps: 1) exons associated with transcripts annotated as “retained_intron” and “read_through” were excluded; 2) exon intervals overlapping within a gene were merged; 3) the intersections of exon intervals overlapping between genes were excluded; 4) the remaining exon intervals were mapped to their respective gene identifier and stored in GTF format. Read counts and TPM values were produced with RNA-SeQC v2.4.2^1^, using the following read-level filters: 1) reads were uniquely mapped (corresponding to a mapping quality of 255 for STAR BAMs); 2 reads were aligned in proper pairs; 3) read alignment distance was <=6 (i.e., alignments must not contain more than six non-reference bases); 4) reads were fully contained within exon boundaries. Reads overlapping introns were not counted. RNA-seq expression outliers were identified and excluded using a multidimensional extension of the statistic described by Wright FA et al.^2^. Briefly, for each tissue, read counts from each sample were normalized using size factors calculated with DESeq2 and log-transformed with an offset of 1; genes with a log-transformed value >1 in >10% of samples were selected, and the resulting read counts were centered and unit-normalized. The resulting matrix was then hierarchically clustered (based on average and cosine distance), and a chi2 p-value was calculated based on Mahalanobis distance. Clusters with ≥60% of samples with Bonferroni-corrected p-values <0.05 were marked as outliers, and their samples were excluded. Samples with <10 million mapped reads were removed.

**Expression quantitative trait loci mapping**. Gene expression values for all samples were normalized using the following procedure: 1) Genes were selected based on expression thresholds of >0.1 TPM in at least 20% of samples and ≥6 reads in at least 20% of samples; 2) expression values were normalized between samples using TMM as implemented in edgeR^3^ and; 3) for each gene, expression values were normalized across samples using an inverse normal transformation. Covariates for eQTL (expression quantitative trait loci) mapping included sex, age, and the top 10 genotyping principal components. A set of covariates identified using the Probabilistic Estimation of Expression Residuals (PEER) method^4^, calculated for the normalized expression matrices was also used. For eQTL analyses, expression values were adjusted for 15 PEER factors to improve the number of eGenes discovered. Next, tensorQTL was used for eQTL quantification as described by the developers^5^. Genotype, phenotype (i.e., normalized gene expression values for each participant) and covariates data was extracted for the participants to include in the analysis. TensorQTL was performed on four different modes (i.e. ‘cis’, ‘cis nominal’, ‘cis independent’ and ‘trans’) using the above-mentioned genotype, phenotype and covariates files.
